## Supplemental Figures for "Fine-scale heterogeneity in population density predicts wave dynamics in dengue epidemics"

### Supplemental material

| **Name** | **Value** | **Parameter** |
|---|---|---|
| $\mu$ | 1/(75*365) $days^{-1}$ | mortality rate |
| $\gamma$ | 1/17 $days^{-1}$ | recovery rate |
| $\beta_0$ | 1.15/17 $days^{-1}$ | mean transmission rate |
| $\omega$ | 2*π/365 $days^{-1}$ | annual frequency of seasonality |
| $\phi_{stochastic}$ | 1.60 | seasonality phase in the stochastic model |
| $\phi_{deterministic}$ | 2.0 | seasonality phase in the deterministic model |
| $\delta_{stochastic}$ | 0.2 | Amplitude of transmission rate in the stochastic model |
| $\delta_{deterministic}$ | 0.99 | Amplitude of transmission rate in the deterministic model |

**Table S1**: Parameters of the *SIR* model (from (*25*))

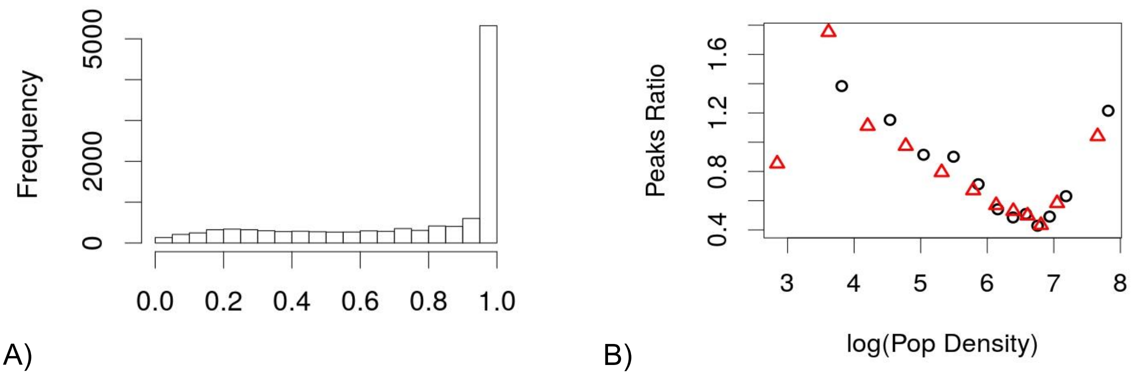


**Fig S1**. **(A)** Histogram of the ratio of unit areas computed with and without non-urban areas. **(B)** Peak ratio vs the natural logarithm of population density. Black circles correspond to population density computed as population over the effective area of the unit, and red triangles correspond to population per unit.

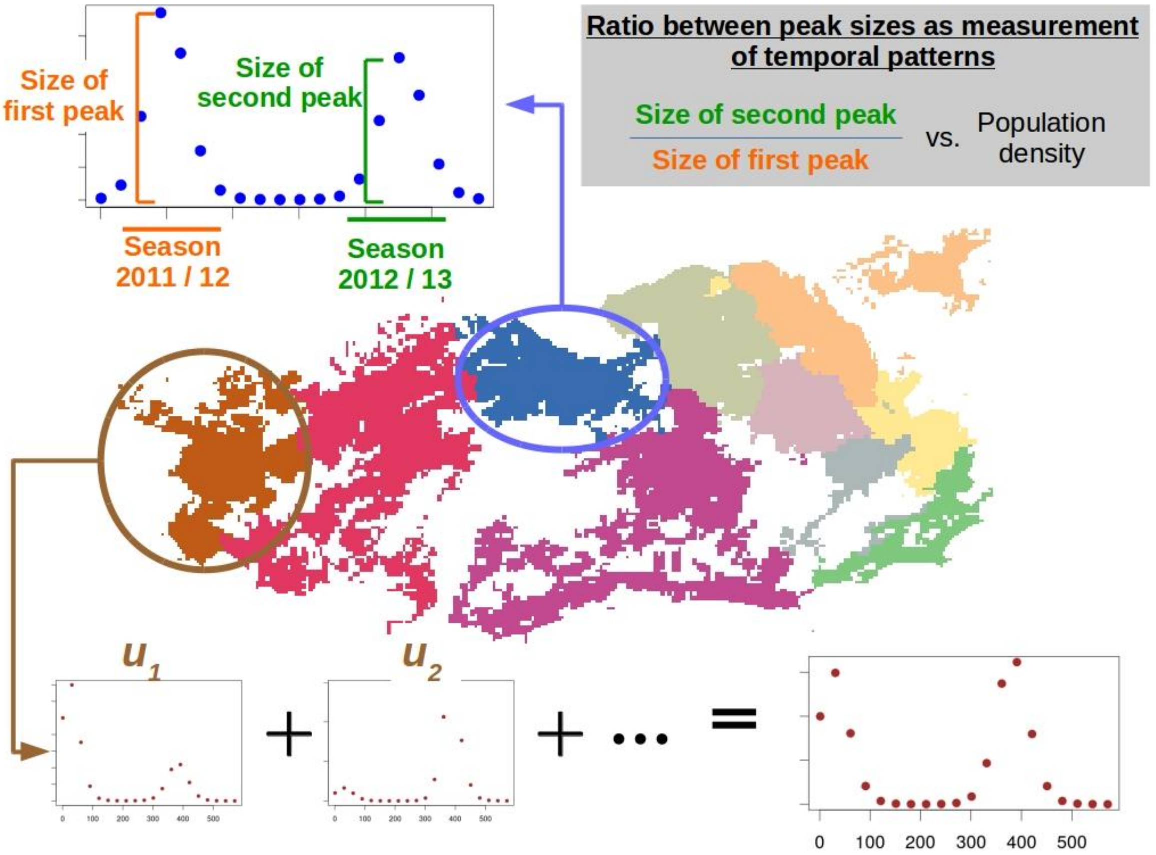


**Fig S2**. Diagram illustrating the aggregation of units when considering incidence patterns. This example considers the 10 administrative regions of Rio de Janeiro and shows the peak ratio for the resulting global time series.

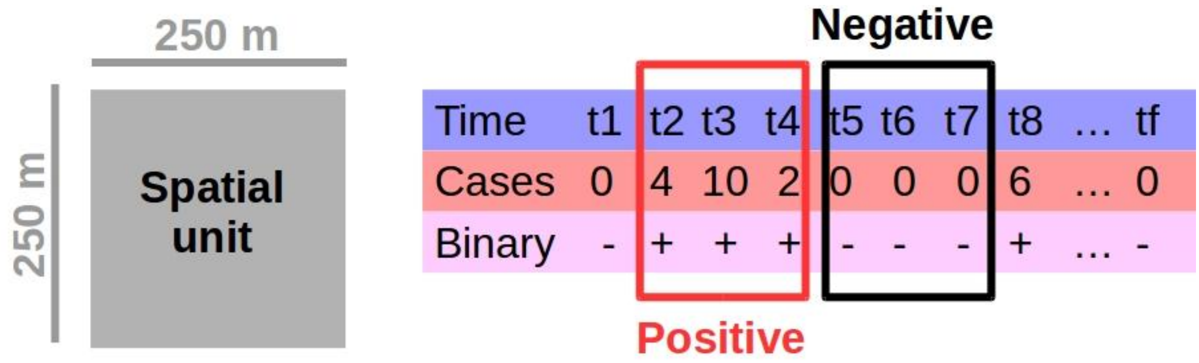


**Fig S3:** Example of a time series of reported cases, illustrating the definition of negative and positive states for the spatial unit.

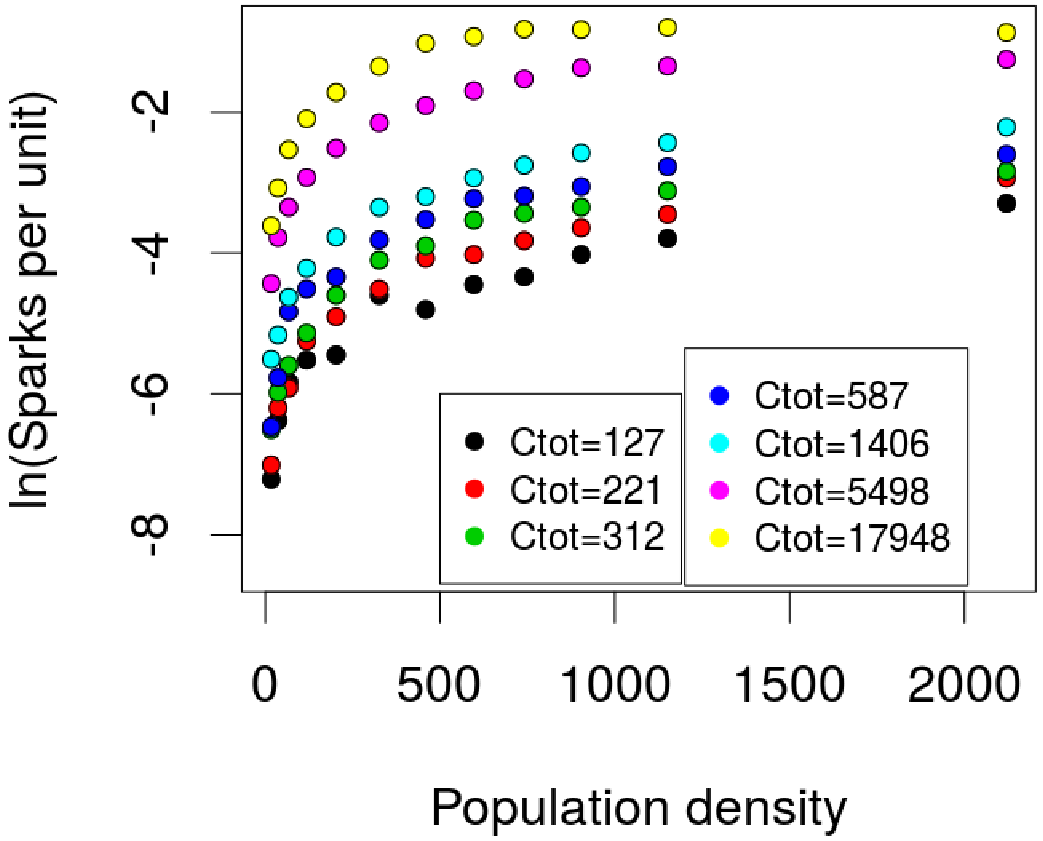


**Fig S4**: Natural logarithm of the number of dengue sparks per spatial unit estimated from the reported data, as a function of the unit's mean population density in Rio de Janeiro city. The colors represent a different total number of cases in the city. As Ctot increases, more sparks are produced.

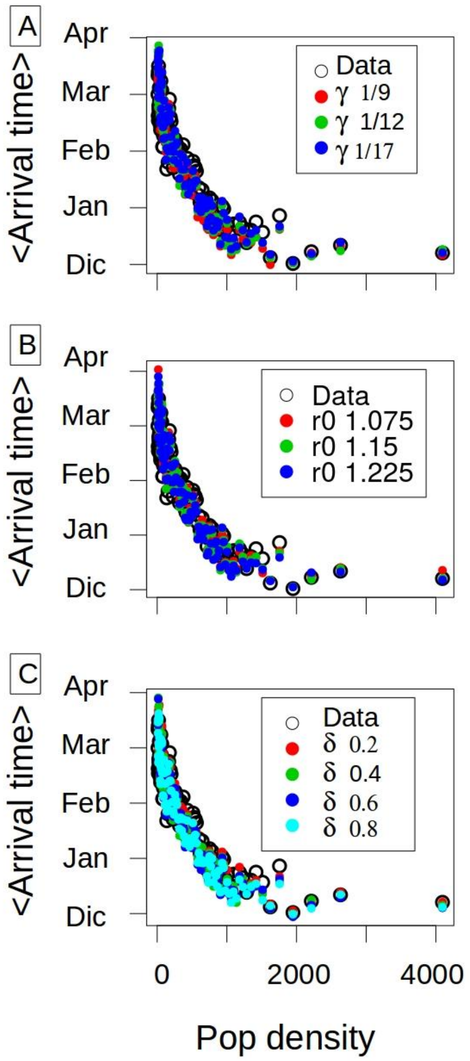


**Fig S5**. Mean arrival time vs population density: **(A)** for different values of $\gamma$ ($\delta$=0.2, r0=1.15, $\rho$ =0.5); **(B)** for different values of r0 ($\delta$=0.2, $\gamma$ =1/17, $\rho$=0.5); and **(C)** for different values of $\delta$ (r0=1.15, $\gamma$=1/17 and $\rho$=0.5).

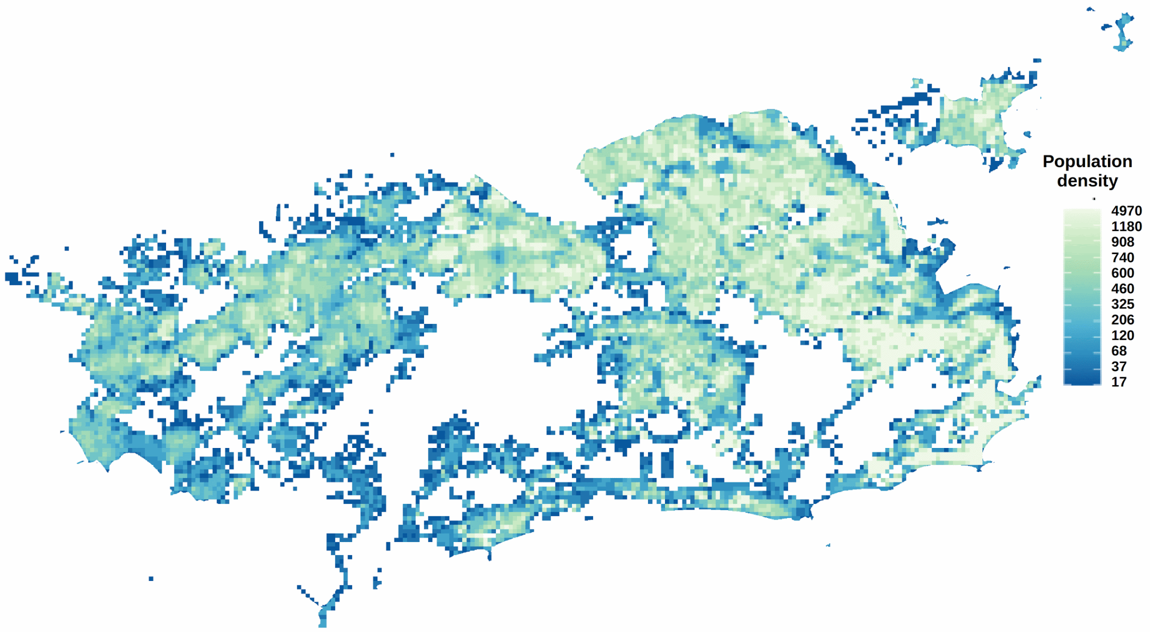


**Fig S6.** Population density computed at the finest resolution of our units.
